# Sex Differences in Non-Acute Myocardial Infarction Cardiogenic Shock: Insights from the Northwell-Shock Registry

**DOI:** 10.1101/2024.12.11.24318890

**Authors:** Shuojohn Li, Abduljabar Adi, Marcy Miller, Fouad Sakr, Jack Jnani, Emily A. Rodriguez, Samuel Tan, Moein Bayat Mokhtari, Ramsis Ramsis, Rebecca Smoller, Gerin R. Stevens, Jaime Hernandez-Montfort, Matthew Griffin, Matthew Pierce, Miguel Alvarez Villela

## Abstract

2.

**Background:** Previous studies had explored the role of patients’ sex in defining clinical presentation, management, and outcomes among patients with acute myocardial infarction (AMI) related cardiogenic shock (CS). However, the effect of sex among patients with non-acute myocardial infarction related CS (nonAMI-CS) is less well defined.

**Method:** Adult patients treated for CS (International Classification of Diseases, Tenth Revision, [ICD-10] code R57.0) between 2016 and 2022 across eleven hospitals within our health system, all utilizing a shared electronic medical record, were included. CS etiologies were defined as AMI, nonAMI and others. For nonAMI-CS patients, stratification by sex was performed to compare the incidence of specific etiologies, baseline characteristics, management and outcomes between men and women. Comparisons were conducted using t-tests, Wilcoxon rank-sum tests or chi-square tests. Logistic regression models were developed to examine the effect of sex on in-hospital mortality, management strategies, and mortality predictors.

**Result:** 2,256 patients admitted for nonAMI-CS were identified. Women comprised 36% of the study cohort and were older and had more medical comorbidities. There was no significant difference in presenting CS severity as measured by Society for Cardiovascular Angiography and Interventions (SCAI) stages, APACHE scores or in-hospital mortality. There were significant differences in the specific etiologies of nonAMI-CS, where women had more valvular cardiogenic shock. Women received fewer invasive interventions including pulmonary artery catheter (PAC) and intra-aortic balloon pump (IABP) when compared to men but similar rates of Impella and venoarterial extracorporeal membrane oxygenation (VA-ECMO). Lastly, women were more likely to be discharged to skilled nursing facilities while men more likely to home.

**Conclusion:** Significant sex-specific differences exist in nonAMI-CS including specific etiologies, management strategies, and factors associated with mortality. Despite older age, more comorbidities and lower rates of invasive management procedures, women had similar survival to men but higher need for skilled facilities upon discharge.

## 3. Background

Cardiogenic shock (CS) is a complex syndrome defined by severe impairment of cardiac function leading to reduced cardiac output, tissue hypoperfusion, and multi-organ dysfunction [1]. Hospital mortality remains high for CS patients despite advancements in percutaneous coronary interventions (PCI) for patients with acute myocardial infarction (AMI) and increased utilization of mechanical circulatory support (MCS) [2–3]. The diversity in etiologies and the variation in disease severity at the time of presentation have limited the development of uniformly effective therapies [4].

There is a growing understanding that patients’ sex plays an important role in defining clinical presentation and outcomes among patients with acute myocardial infarction related cardiogenic shock (AMI-CS) [5–6]. Whether this effect of sex also plays a role among patients with non-AMI related CS (nonAMI-CS), is less well described.

This knowledge gap is particularly important because nonAMI-CS includes a more heterogeneous patient population and represents a majority of cases in contemporary CS registries [7–8]. Understanding the sex-related differences existing in this group can help better describe the epidemiology of CS as a whole contributing to the development of sex-specific management strategies and improve clinical trial design.

In this study, we describe the clinical differences between men and women in a population of patients with a variety of nonAMI-CS managed within a multi-level of care metropolitan health system.

## 4. Methods

The Northwell-Shock Registry is a multi-center retrospective observational study of patients with CS. It was approved by the Northwell Health Institutional Review Board. This registry was deemed to pose minimal risk to the study subjects and a waiver of informed consent was obtained. Patient data are de-identified and stored in a secure password-protected database.

### 4.1 Study population

The characteristics of our registry have been previously described [9]. Briefly, it is composed of data from 11 Northwell Health hospitals, including 4 cardiac surgery sites within our 23-hospital network, in the New York metropolitan area. All participating sites share a common electronic health record (Sunrise Clinical Manager). All patients over 18 years of age discharged from these sites between January 2016 and August 2022 with a principal or secondary diagnosis of CS are included. CS was identified using the International Classification of Disease (ICD)-10 code R57.0.

Data were collected for each patient directly from the chart including demographic information, comorbidities, laboratory results, level of care at discharge, treatments provided, complications, and hospital outcomes using ICD-10 procedural and diagnostic codes (Supplemental Table S1 & S2). Comorbidities were summarized using ICD-10 version of the Charlson Comorbidities Index (CCI) [10]. Baseline vital signs and laboratory values were defined as the first available laboratory value results within 24 hours of hospital presentation. Baseline pulmonary artery catheter derived hemodynamics were defined as the first available in the medical record.

Hospital levels of care for CS were classified according to MCS capabilities [9]. When analyzing adverse outcomes, advanced MCS was defined as Venoarterial extracorporeal membrane oxygenation (Maquet Cardiopulmonary AG, Hirrlingen, Germany or CentriMag; Abbott Laboratories, Abbott Park, IL) and/or Impella devices (Abiomed, Danvers, MA). The vasoactive-inotropic score (VIS) was calculated based on the number and dosage of inotropes and vasopressors at admission (VIS 0h) and at the time of maximal dosing (VIS max) [11]. Major complications included major bleeding defined using the Bleeding Academic Research Consortium (BARC) scale, arterial thrombosis requiring surgical intervention, new renal replacement therapy and sepsis defined as clinical worsening with a documented or suspected infection. Information regarding these complications was extracted manually from the charts.

### 4.2 Shock etiologies and Severity of Shock

The primary etiology of CS was divided into three major categories using a modification of the classification proposed by the Shock Academic Research Consortium (SHARC) [12] based on the principal discharge diagnoses codes associated with CS: (1) acute myocardial infarction (AMI) including ST-elevation myocardial infarction (STEMI) and non-ST-elevation myocardial infarction (NSTEMI); (2) NonAMI-CS including “de novo” and acute on chronic heart failure (HF) as well as “secondary CS” due to arrhythmias, infective endocarditis and valvular disease; (3) Other etiologies including post-cardiotomy CS, pericardial disorders, mixed septic-cardiogenic shock, pulmonary embolism as well as other non-cardiac causes which have been previously described [9]. For this analysis, only patients within the nonAMI-CS category were included (Supplemental Figure S1).

Specific sub-categories are described within the valvular and arrhythmic etiologies. A complete list of ICD-10 codes assigned to each etiologic group is provided in the supplemental material (Supplemental Table S1 & S2).

The severity of CS is described according to the Society for Cardiovascular Angiography and Interventions (SCAI) stages classification using the objective criteria previously described by the cardiogenic shock working group (SCAI-CSWG) [13–14]. The APACHE IV score at the time of intensive care unit (ICU) admission was calculated for patients admitted to an ICU at any point during their hospital stay [15].

### 4.3 Statistical analysis

Data analysis was conducted using RStudio version 4.4.1, while graphs were generated using GraphPad Prism version 10.3.1. Missing values were handled through multiple imputation via chained equation (using the ‘mice’ R package). Continuous variables were reported as mean ± SD for normally distributed data, or as median (25^th^ percentile, 75^th^ percentile) for non-normally distributed data. Categorical variables were summarized as frequencies and percentages.

Comparison of continuous variables were performed using t-test for normally distributed data and Wilcoxon rank-sum test for non-normally distributed data. Categorical variables were compared using chi-square tests.

To examine differences in management strategies by sex, a multivariate logistic regression model was developed with sex (women) as the outcome and interventions as predictors. Additionally, the effect of sex on in-hospital mortality was examined via multivariate adjusted analysis. Finally, multivariate logistic regression models were constructed separately for men and women, using in-hospital mortality as the outcome to explore sex-specific predictors of mortality.

The primary outcome was in-hospital mortality. Secondary outcomes were in-hospital mortality stratified by specific etiology of CS, disposition at hospital discharge for survivors and rate of major complications in patients receiving advanced MCS (e.g. Impella and/or Venoarterial extracorporeal membrane oxygenation).

## 5. Results

### 5.1 Baseline characteristics

A total of 2,256 patients were included in this analysis (Supplemental Figure S1), with women comprising 36% of the cohort. Women were older (71.3 vs. 67.7 year; p < 0.01), had smaller body surface area (1.84 vs. 2.06 m^2^; p < 0.01) and more comorbidities based on Charlson Comorbidities Index (CCI) (6.7 vs. 6.3; p = 0.02) than men [10]. Women had a lower prevalence of previous coronary artery disease (33% vs. 47%; p < 0.01) and chronic kidney disease (CKD) (40% vs. 46%; p < 0.01) but lower glomerular filtration rate on presentation (41.2 vs. 45.2 mL/min/1.73m^2^; p < 0.01). Previous history of heart failure was highly prevalent in both sexes (women 74% vs. Men 77%; p = 0.11) (Table 1).

**Table 1.** Baseline demographics, comorbidities, admission characteristics and laboratory data. *ALT = alanine transaminase, BMI = body mass index, BSA = body surface area, CCI = Charlson Comorbidity Index, CI = cardiac index, CO = cardiac output, CVP = central venous pressure, ECMO = extracorporeal membrane oxygenation, EF = ejection fraction, eGFR = estimated glomerular filtration rate, HT = heart transplant, IABP = intra-aortic balloon pump, LVEDD = left ventricular end-diastolic diameter, LVAD = durable left ventricular assist device, MAP = mean arterial pressure, mPAP = mean pulmonary artery pressure, MCS = mechanical circulatory support, PAD = pulmonary artery diastolic pressure, PAS = pulmonary artery systolic pressure, pVAD = percutaneous ventricular assist device, SCAI = Society of Cardiovascular Angiography and Interventions, WBC = white blood cell*

| Parameter | Women<br>n = 803 | Men<br>n = 1453 | p value |
| --- | --- | --- | --- |
| Age (year) | 71.3 ± 14.6 | 67.7 ± 13.8 | **<0.01** |
| BMI (kg/m<sup>2</sup>) | 30.1 ± 10.1 | 29.6 ± 9.7 | 0.22 |
| BSA (m<sup>2</sup>) | 1.84 ± 0.33 | 2.06 ± 0.33 | **<0.01** |
| Race (%) |  |  | 0.20 |
| White | 403 (50) | 760 (52) |  |
| Black/African American | 199 (25) | 310 (21) |  |
| Asian | 54 (7) | 119 (8) |  |
| Other | 147 (18) | 264 (18) |  |
| Comorbidities (%) |  |  |  |
| Diabetes mellitus | 322 (40) | 624 (43) | 0.21 |
| Hypertension | 551 (69) | 976 (67) | 0.51 |
| Chronic kidney disease | 322 (40) | 673 (46) | **<0.01** |
| Heart failure | 595 (74) | 1122 (77) | 0.11 |
| Coronary artery disease | 264 (33) | 681 (47) | **<0.01** |
| Atrial fibrillation | 403 (50) | 742 (51) | 0.72 |
| CCI | 6.7 ± 3.66 | 6.3 ± 3.66 | **0.02** |
| Cardiac arrest at admission | 18 (2) | 45 (3) | 0.30 |
| Level of care |  |  | 0.71 |
| LVAD and HT center | 345 (43) | 626 (43) |  |
| ECMO center | 383 (48) | 706 (49) |  |
| IABP and pVAD center | 20 (2) | 39 (3) |  |
| No MCS capability | 55 (7) | 82 (6) |  |
| Transfer | 48 (6) | 129 (9) | **0.02** |
| Admission APACHE score | 76.1 ± 32.4 | 74.2 ± 32.1 | 0.19 |
| Admission SCAI stage (%) |  |  | 0.27 |
| A | 256 (32) | 442 (30) |  |
| B | 162 (20) | 338 (23) |  |
| C | 94 (12) | 190 (13) |  |
| D | 111 (14) | 194 (13) |  |
| E | 147 (18) | 232 (16) |  |
| Unrecorded | 33 (4) | 57 (4) |  |
| EF (%) (n = 346 vs. 657) | 30 (20, 55) | 23 (15, 35) | **<0.01** |
| LVEDD (cm) (n = 348 vs. 705) | 4.8 (4.2, 5.6) | 5.8 (5.0, 6.4) | **<0.01** |
| Hemodynamics |  |  |  |
| Heart rate (BPM) | 84 (70, 100) | 86 (72, 102) | **0.01** |
| MAP (mmHg) | 68 (59, 78) | 71 (61, 80) | **<0.01** |
| RAP (n = 220 vs. 515) | 12 (7, 17) | 12 (8, 17) | 0.40 |
| PAS (n = 220 vs. 515) | 44 (34, 57) | 47 (37, 60) | **0.03** |
| PAD (n = 220 vs. 515) | 22 (17, 28) | 23 (17, 31) | 0.09 |
| mPAP (n = 220 vs. 515) | 30 (22, 37) | 31 (24, 41) | **0.05** |
| CO (n = 220 vs. 515) | 4 (3.1, 5) | 4.2 (3.3, 5.3) | **0.03** |
| CI (n = 220 vs. 515) | 2.1 (1.7, 2.6) | 2.1 (1.7, 2.7) | 0.47 |
| Laboratory |  |  |  |
| Lactate (mmol/L) | 2.2 (1.7, 3.8) | 2.2 (1.7, 3.8) | 0.71 |
| Creatinine (mg/dL) | 1.55 (1.02, 2.53) | 1.82 (1.28, 2.87) | **<0.01** |
| eGFR (mL/min/1.73m<sup>2</sup>) | 41.2 (23.5, 69.0) | 45.2 (27.6, 69.6) | **<0.01** |
| ALT (IU/L) | 28 (16, 66) | 33 (19, 87) | **<0.01** |
| Bilirubin (mg/dL) | 0.8 (0.5, 1.4) | 1.0 (0.6, 1.8) | **<0.01** |
| Sodium (mEq/L) | 137 (133, 140) | 137 (133, 139) | 0.24 |
| Hemoglobin (g/dL) | 11.1 (9.6, 12.5) | 12.1 (10.2, 13.6) | **<0.01** |
| WBC (k/uL) | 9.3 (6.4, 13.3) | 8.8 (6.6, 12.3) | 0.16 |
| Platelet (k/uL) | 189 (133, 246) | 177 (126, 234) | **<0.01** |
| Albumin (g/dL) | 3.4 (3, 3.8) | 3.4 (3.1, 3.8) | 0.1 |

On transthoracic echocardiogram, women had higher left ventricular (LV) ejection fraction (30% vs. 23%; p < 0.01) and smaller LV end-diastolic diameter (4.8 vs. 5.8 cm; p < 0.01).

Hemodynamically, women showed slower heart rate and lower mean arterial pressure at presentation. Invasive pulmonary artery catheter (PAC) derived hemodynamics showed that women also had lower pulmonary artery systolic and mean pressures with similar central venous pressures and cardiac index (Table 1).

Notable differences in laboratory analyses were lower alanine transferase (ALT), total bilirubin, hemoglobin and higher platelet count in women. Lactate levels at presentation were similar in both sexes (Table 1).

### 5.2 CS Etiologies and Severity

Significant differences were found in the specific etiologies leading to nonAMI-CS (Table 2). Similar prevalences of heart failure related CS (HF-CS) were noted between women and men (59% women vs. 63% men, p = 0.11). However, women had lower incidence of acute on chronic HF (51% vs. 56%, p = 0.04) and similar rates of de novo HF (8% vs. 7%, p = 0.39), when compared to men (Figure 1).

**Figure 1.**
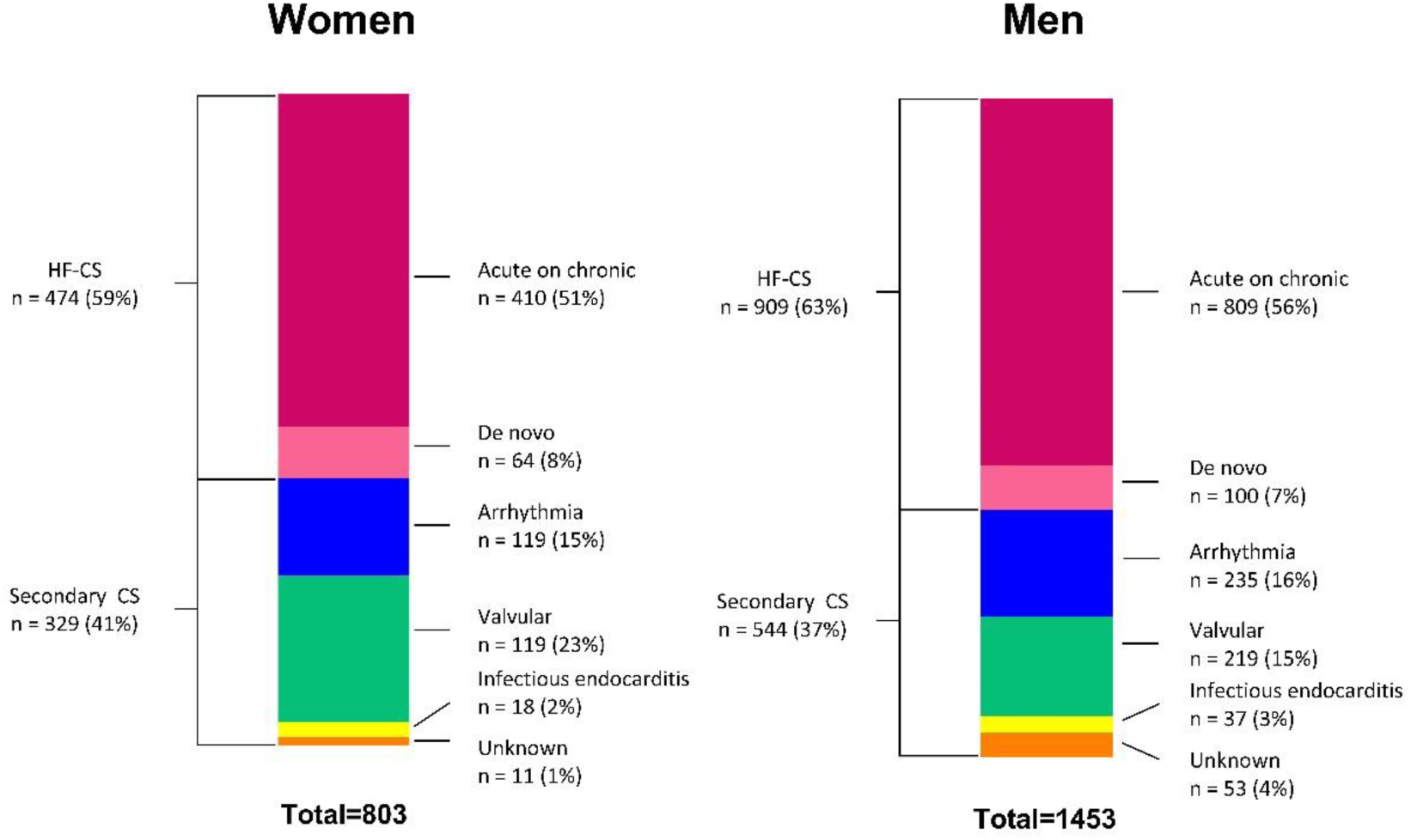
Etiologies of non-acute myocardial infarction related cardiogenic shock by patient sex. *CS = Cardiogenic shock, HF-CS = heart failure related cardiogenic shock*.

**Table 2.** Etiologies of Non-Acute Myocardial Infarction Cardiogenic Shock. *CS = Cardiogenic shock*

| Etiology (%) | Women | Men | p value |
| --- | --- | --- | --- |
|  | n =803 | n = 1453 |  |
| Heart failure-CS | 474 (59) | 909 (63) | 0.11 |
| Acute on chronic | 410 (51) | 809 (56) | **0.04** |
| De novo | 64 (8) | 100 (7) | 0.39 |
| Secondary CS | 329 (41) | 544 (37) |  |
| Arrhythmia | 119 (15) | 235 (16) | 0.43 |
| Atrial | 38 (32) | 63 (27) | 0.38 |
| Ventricular | 29 (24) | 121 (51) | **<0.01** |
| Bradyarrhythmia | 50 (42) | 47 (20) | **<0.01** |
| Unrecorded | 2 (2) | 4 (2) |  |
| Valvular | 181 (23) | 219 (15) | **<0.01** |
| Aortic stenosis | 38 (21) | 65 (30) | 0.10 |
| Aortic regurgitation | 10 (6) | 37 (17) | **<0.01** |
| Mitral stenosis | 5 (3) | 2 (1) | 0.31 |
| Mitral regurgitation | 74 (41) | 52 (24) | **<0.01** |
| Prosthetic valve | 7 (4) | 24 (11) | **0.02** |
| Right sided valve | 5 (3) | 5 (2) | 1.0 |
| Multivalvular | 42 (23) | 31 (14) | **0.03** |
| Unrecorded | 0 (0) | 3 (1) |  |
| Infectious endocarditis | 18 (2) | 37 (3) | 0.76 |
| Unknown etiologies | 11 (1) | 53 (4) |  |

Women more frequently had valvular related CS compared to men (23% vs. 15%; p < 0.01) (Figure 1) and within the valvular CS group, women were more likely to have mitral regurgitation (41% vs. 24%; p < 0.01) and multivalvular disease (23% vs. 14%; p = 0.03), but less likely to have aortic regurgitation (6% vs. 17%; p < 0.01) or prosthetic valve disease (4% vs. 11%; p = 0.02) as a cause for CS. Aortic stenosis related CS was similarly prevalent in men and women (21% vs. 30%; p=0.1) (Table 2).

Arrhythmia as a trigger of CS was equally frequent in both sexes (15% vs. 16%; p = 0.43) However, women were more affected by bradyarrhythmia (42% vs. 20%; p < 0.01) and men were more affected by ventricular arrythmias (24% vs. 51%; p < 0.01) (Table 2).

CS severity was measured using SCAI-CSWG stages [13–14] and APACHE score on ICU admission [15]. No significant sex-based differences were seen in initial SCAI stage distribution (p = 0.27) or average APACHE scores (p = 0.19). Similarly, there was no difference in maximal SCAI stages reached during the CS hospitalization (p = 0.48) (Table 1 & Supplemental Figure S3).

### 5.3 Management strategies

Women had lower rates of PAC use for invasive hemodynamic monitoring (44% vs. 53%; p < 0.01) and MCS devices (15% vs. 22%; p < 0.01) driven mostly by lower use of intra-aortic balloon pump (IABP) while venoarterial extracorporeal membrane oxygenation (VA-ECMO) and Impella were used with equal frequency in both sexes (Table 3 & Supplemental Figure S4). Among patients who received Impella, device types used were 2.5 in 4%, CP in 76%, 5.0 in 7%, 5.5 in 3%, RP in 3%, unknown in 7%.

**Table 3.** Non-acute myocardial infarction related cardiogenic shock management. *CABG = Coronary artery bypass graft, IABP = intra-aortic balloon pump, LVAD = durable left ventricular assist device, MCS = mechanical circulatory support, NIPPV = noninvasive positive pressure ventilation, PCI = percutaneous coronary intervention, RRT = renal replacement therapy, VA-ECMO = venoarterial extracorporeal membrane oxygenation, VIS = vasoactive-inotropic score*

| Intervention (%) | Women<br>n = 803 | Men<br>n = 1453 | p value |
| --- | --- | --- | --- |
| Catecholamine | 287 (36) | 515 (35) | 0.92 |
| Epinephrine | 101 (13) | 194 (13) | 0.65 |
| Norepinephrine | 287 (36) | 515 (35) | 0.92 |
| Inotrope | 322 (40) | 608 (42) | 0.45 |
| Dobutamine | 234 (29) | 435 (30) | 0.72 |
| Milrinone | 188 (23) | 405 (28) | **0.02** |
| Dopamine | 32 (4) | 35 (2) | **0.05** |
| VIS score at 0h<br>(n = 189 vs. 323) | 10 (5, 25) | 8 (4, 24) | 0.19 |
| VIS score at max<br>(n = 366 vs. 623) | 21 (9, 65) | 22 (8, 52) | 0.44 |
| MCS | 117 (15) | 322 (22) | **<0.01** |
| IABP | 80 (68) | 215 (67) |  |
| Impella | 18 (15) | 55 (17) |  |
| VA-ECMO | 18 (15) | 45 (14) |  |
| Impella + VA-ECMO | 10 (9) | 17 (5) |  |
| LVAD | 12 (1) | 54 (4) | **<0.01** |
| Transplant | 14 (2) | 22 (2) | 0.81 |
| Pulmonary artery catheter | 353 (44) | 773 (53) | **<0.01** |
| Left heart catheterization | 191 (24) | 385 (26) | 0.17 |
| PCI | 23 (3) | 49 (3) | 0.59 |
| CABG | 29 (4) | 93 (6) | **<0.01** |
| Valvular procedure | 172 (21) | 258 (18) | **0.04** |
| Surgical | 117 (68) | 155 (60) |  |
| Percutaneous | 39 (23) | 42 (16) |  |
| Unrecorded | 16 (9) | 61 (24) |  |
| Blood transfusion | 277 (34) | 406 (28) | **<0.01** |
| Mechanical ventilation | 272 (34) | 499 (34) | 0.86 |
| NIPPV | 180 (22) | 355 (24) | 0.30 |
| RRT | 107 (13) | 240 (17) | **0.05** |

Regarding other ICU therapies, women received less renal replacement therapy (RRT), more blood transfusions, and similar rates of mechanical ventilation. While no differences were seen in the overall use of vasopressors (p = 0.92) or inotropes (p = 0.45), nor in initial or maximal vasoactive-inotropic score (VIS), there was a slightly higher use of Milrinone in men (28% vs 23%; p<0.01) and of Dopamine in women (4% vs. 2%; n = 0.05) (Table 3).

Coronary artery bypass graft surgery (CABG) occurred less often in women compared to men (4% vs. 6%, p < 0.01), while percutaneous and surgical valvular interventions were more frequent among women (21% vs. 18%; p=0.04) (Table 3). Other invasive management procedures such as left heart catheterization (LHC) and PCI were used equally in both sexes.

The overall use of heart replacement therapies was low, but the frequency of durable left ventricular assist device (LVAD) implantation was lower in women than in men (1% vs. 4%; p<0.01) while heart transplantation was used at similar rates in both sexes (2% vs. 2%) (Table 3).

After adjusting for age, BSA, baseline lactate and eGFR, and history of hypertension or diabetes, blood transfusion (OR = 1.63, 95% CI: 1.29-2.07) was the only intervention performed more frequently in women, while IABP (OR = 0.65, 95% CI: 0.47-0.89), PAC (OR = 0.77, 95% CI: 0.62-0.95), CABG (OR = 0.43, 95% CI: 0.26-0.68) and durable LVAD implantation (OR = 0.31, 95% CI: 0.15-0.63) were all less frequent in women (Supplemental Figure S5).

### 5.4 Complications in Advanced Mechanical Circulatory Support

To evaluate the safety of advanced MCS devices defined as Impella, VA-ECMO or the combination of the two, we analyzed the adverse events occurring in men and women treated with these MCS types.

Use of these devices was low overall and similar in both sexes (Women=46; 6% vs. Men=117, 8%). No differences were found in the rates of major bleeding (BARC 3-5), arterial thrombosis requiring surgery, need for renal replacement therapy (RRT) or sepsis in this subgroup (Table 4).

**Table 4.** Major complications in non-acute myocardial infarction associated cardiogenic shock patients treated with advanced mechanical circulatory support. *BARC = Bleeding Academic Research Consortium, MCS = mechanical circulatory support, RRT = renal replacement therapy, VA-ECMO = venoarterial extracorporeal membrane oxygenation*.

| Advanced MCS | Women | Men | p value |
| --- | --- | --- | --- |
| complications (%) | n = 46 | n = 117 |  |
| **Impella only** | **n = 18** | **n = 55** |  |
| BARC 3-5 bleeding | 1 (6) | 4 (7) | 1.0 |
| Arterial thrombosis | 1 (6) | 2 (4) | 1.0 |
| Transfusion | 4 (22) | 9 (16) | 0.83 |
| RRT | 7 (56) | 13 (24) | 0.34 |
| Sepsis | 1 (6) | 7 (13) | 0.68 |
| **VA-ECMO only** | **n = 18** | **n = 45** |  |
| BARC 3-5 bleeding | 0 (0) | 4 (9) | 0.46 |
| Arterial thrombosis | 0 (0) | 0 (0) | NA |
| Transfusion | 1 (6) | 11 (24) | 0.46 |
| RRT | 2 (11) | 6 (13) | 1.0 |
| Sepsis | 1 (6) | 2 (4) | 1.0 |
| **Impella and VA-ECMO** | **n = 10** | **n = 17** |  |
| BARC 3-5 bleeding | 3 (30) | 3 (18) | 0.79 |
| Arterial thrombosis | 1 (10) | 1 (6) | 1.0 |
| Transfusion | 1 (10) | 6 (35) | 0.32 |
| RRT | 3 (30) | 3 (18) | 0.79 |
| Sepsis | 2 (20) | 2 (12) | 0.98 |

### 5.5 Hospital Outcomes

Men and women had similar in-hospital mortality (27% vs. 25%; p = 0.21) and length of hospital stay (12 vs. 12 days; p = 0.39). (Table 5). However, women were less likely to be discharged to home compared to men (14% vs. 20%) and more likely to be discharged to rehabilitation, skilled nursing, or hospice facilities (p < 0.01).

**Table 5.** Hospital outcome of non-acute myocardial infarction related cardiogenic shock. *HF = heart failure*

| Outcome | Women | Men | p value |
| --- | --- | --- | --- |

|  | n =803 | n = 1453 |  |
| --- | --- | --- | --- |
| Disposition at discharge (%) |  |  | <.01 |
| Home | 110 (14) | 290 (20) |  |
| Rehab or skilled facility | 379 (47) | 644 (44) |  |
| Hospice | 44 (6) | 45 (3) |  |
| Unrecorded | 50 (6) | 112 (8) |  |
| Length of stay (day) | 12 (6, 20) | 12 (6, 21) | 0.39 |
| In-hospital mortality (%) | 220 (27) | 362 (25) | 0.21 |
| Acute on chronic HF | 123 (30) | 200 (25) | 0.06 |
| De novo HF | 32 (50) | 31 (31) | **0.02** |
| Arrhythmia | 30 (25) | 69 (29) | 0.49 |
| Valvular | 25 (14) | 29 (13) | 0.98 |
| Infectious endocarditis (%) | 5 (28) | 14 (38) | 0.66 |

Female sex is not an independent predictor of mortality in multivariate analysis (HR = 1.16, 95% CI = 0.68-1.96). However, multivariate logistic regression model constructed separately for women and men revealed different mortality predictors between sexes. Significant multivariable factors associated with higher mortality were a history of hypertension in women (OR = 1.84, 95% CI = 1.19-2.89), while higher BSA (OR = 1.16, 95% CI = 1.01-1.32) and treatment with vasopressors (OR = 1.46, 95% CI = 1.01-2.12) were associated with higher mortality in men.

Factors associated with lower mortality were durable LVAD implantation (OR = 0.22, 95% CI = 0.07-0.68) and treatment with PCI (OR = 0.36, 95% CI = 0.13-0.996) in men, and the use of PAC (OR = 0.55, 95% CI = 0.37-0.86) in women.

Older age, the need for RRT or mechanical ventilation as well as treatment with Impella, were all associated with increased mortality in both sexes (Figure 2). These factors remained as significant predictors of mortality in a multivariate adjusted model excluding CS etiologies as covariates (Supplemental Figure S6)

**Figure 2.**
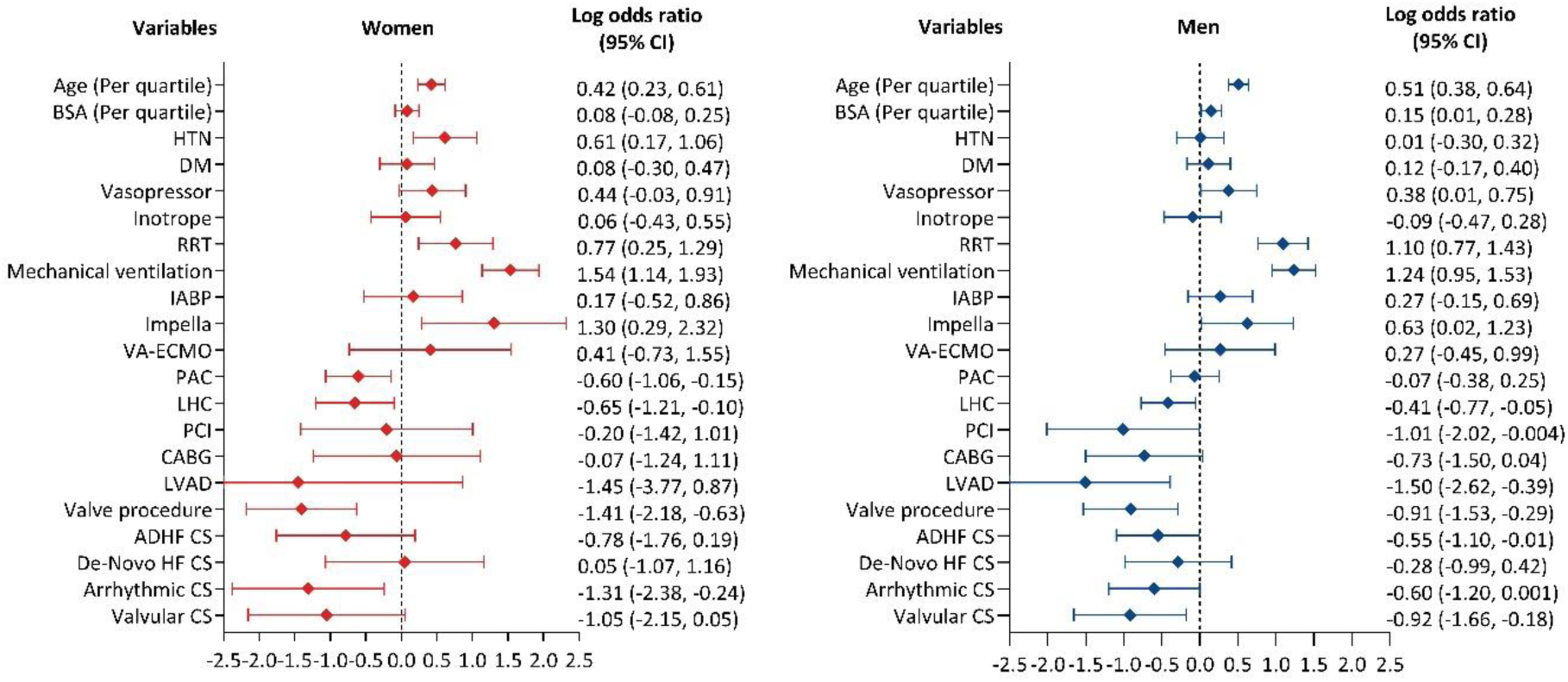
Multivariate logistic regression model of mortality associated factors in women and men treated with non-acute myocardial infarction related cardiogenic shock. *ADHF = acute decompensated heart failure, BSA = Body surface area, CABG = coronary artery bypass graft, CS = cardiogenic shock, DM = Diabetes mellitus, HF = heart failure, HTN = hypertension, IABP = intra-aortic balloon pump, LHC = left heart catheterization, LVAD = left ventricular assist device, PAC = pulmonary artery catheter, PCI = percutaneous coronary intervention, RRT = renal replacement therapy, VA-ECMO = venoarterial extracorporeal membrane oxygenation*.

## 5. Discussion

This study examined the sex-related differences in patients with nonAMI-CS across a large, diverse cohort of patient treated within a multi-tier healthcare system in the New York metropolitan area. Our main findings are summarized in the Central Figure (Figure 3).

**Figure 3.**
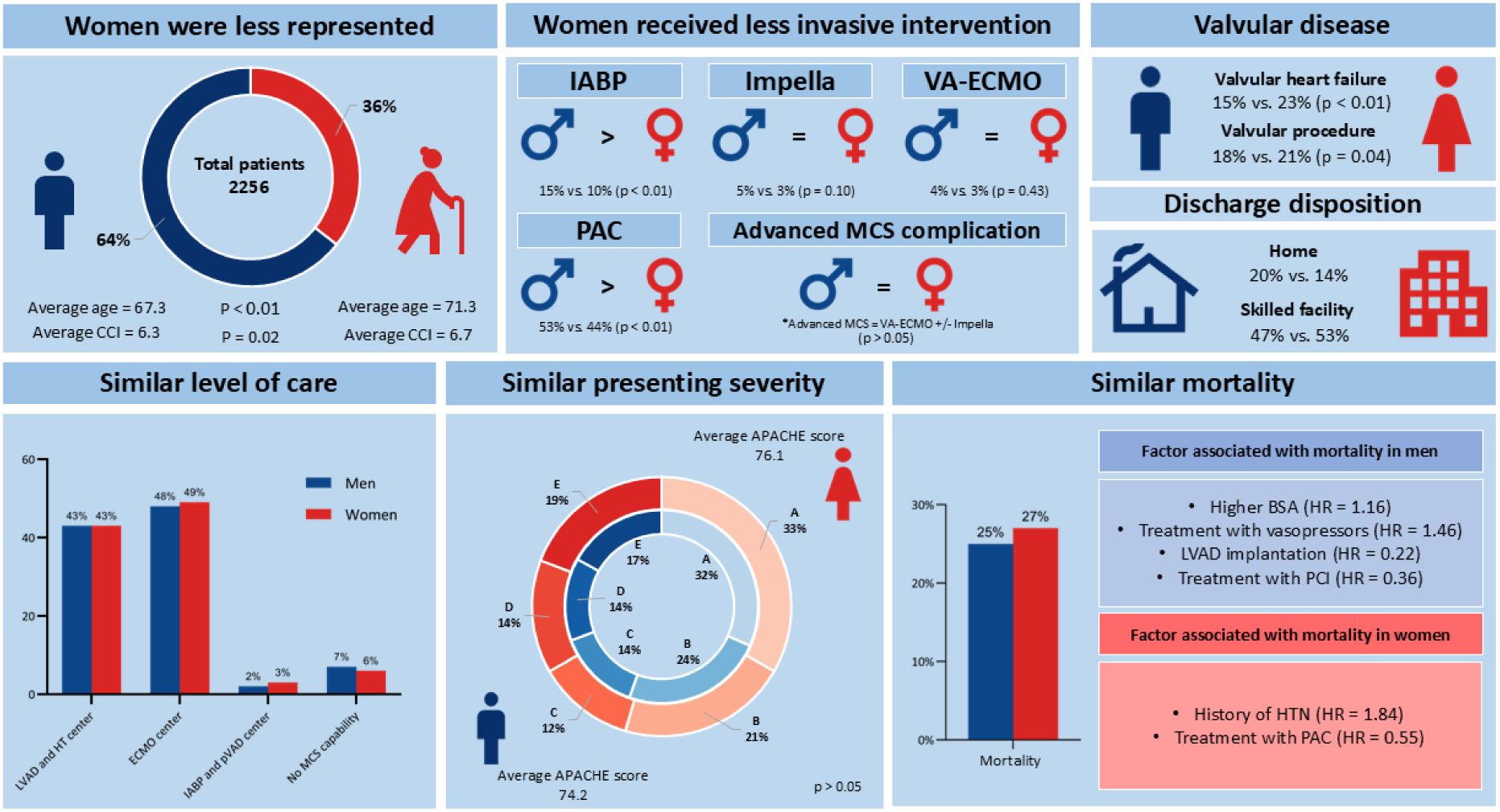
Central illustration. *BSA = body surface area, CCI = Charlson Comorbidities Index, HTN = hypertension, IABP = intra-aortic balloon pump, LVAD = left ventricular assist device, MCS = mechanical circulatory support, PCI = percutaneous coronary intervention, PAC = pulmonary artery catheter, pVAD = percutaneous ventricular assist device, VA-ECMO = venoarterial extracorporeal membrane oxygenation*.

Women were a minority in this cohort (36%) and presented with older age and higher comorbidity burden than men. In line with prior studies, they had lower rates of coronary artery disease (CAD) and chronic kidney disease (CKD) and exhibited relatively better preserved left ventricular ejection fraction (LVEF) [16–19]. Despite these differences, the severity of shock at presentation was comparable between men and women, as assessed by SCAI-CSWG stages, APACHE scores, initial lactate levels, and vasoactive-inotropic scores (VIS).

Important sex-based variations in the prevalence of specific nonAMI-CS etiologies were also identified in this study.

Heart failure–related CS (HF-CS) was the most prevalent etiology with similar prevalence in both sexes, affecting 59% of women and 63% of men. In line with previous reports, women had higher prevalence of “de-novo” HF possibly related to the lower prevalence of previous CAD [7]. In multivariable analysis, HF-CS was associated with lower mortality in men but not in women which may be related to the higher prevalence of “de novo” HF in women, an etiology previously associated with poorer clinical outcomes [20].

Valvular-related CS was the most common secondary etiology and was the only etiologic group with different prevalence among the sexes with higher prevalence in women than in men (23% vs. 15%, p<0.01). Within this group, mitral regurgitation and multivalvular disease were more common in women, while aortic regurgitation was more common among men. This distribution is reflective of the overall pattern of valvular heart disease seen in both sexes [21]. A recent study similarly found that patients with valvular CS (VCS) were more often older and female and reported a higher 30-day mortality for those with VCS compared to those without (28% vs.

20%) [22]. This contrasts with our findings where hospital mortality was the lowest in the VCS group for both men (13%) and women (14%), compared to other CS etiologic groups. Notably, in our cohort, approximately 20% of patients underwent transcatheter valve interventions, which reflects the evolving role of percutaneous approaches in VCS management [22].

Arrhythmia-induced CS was the following most common etiology and was also similarly prevalent in both sexes, but with important differences in the types of arrhythmias triggering CS. Men were more likely to experience ventricular arrhythmias, whereas bradyarrhythmias were more frequent in women. These trends align with established epidemiological patterns in arrhythmia prevalence by sex, where ventricular arrhythmias are more common in men [23].

Among women, arrhythmia-related CS was the only etiology independently associated with lower mortality.

Infective endocarditis (IE)-related CS was a less common etiology in this cohort, with a similar prevalence in men and women. While IE-CS was not significantly associated with mortality in our analysis, this group remains an important clinical challenge given its potential for rapid clinical deterioration and the existent prominent uncertainties in management.

In addition to sex-based differences in etiology, this study found notable variations in management strategies. Women received lower adjusted rates of pulmonary artery catheterization (PAC), intra-aortic balloon pump (IABP) support, coronary artery bypass grafting (CABG), and durable left ventricular assist devices (LVAD). However, rates of advanced mechanical circulatory support (MCS) devices such as VA-ECMO and Impella were similar between sexes, with no observed differences in complication rates. These results align with recent findings from the Cardiogenic Shock Working Group (CSWG) and the Critical Care Cardiology Trials Network (CCCTN), which reported less aggressive interventions in women [7–8]. In our study, these lower intervention rates did not negatively impact outcomes, as hospital mortality and complication rates were similar between sexes, even with fewer invasive procedures in women.

Unadjusted hospital mortality was comparable between men and women, and multivariable analysis revealed that sex was not an independent predictor of mortality despite the described baseline differences. However, specific factors associated with mortality differed by sex: in women, hypertension increased mortality risk, whereas in men, larger body surface area and vasopressor use were significant predictors of higher mortality. Shared predictors of poor outcomes included advanced age, mechanical ventilation, and treatment with the Impella device. These findings underscore the need for further research to evaluate device-specific mortality trends in nonAMI-CS, particularly with newer Impella types such as the 5.5 [24].

Finally, discharge disposition showed that women were less likely to return home and more often required discharge to skilled nursing facilities (SNF) or hospice. This difference may reflect the older age and higher comorbidity burden observed in women, underscoring the higher need for post-hospital support for female CS survivors. Prior studies have linked SNF discharge with reduced 30-day readmission in nonAMI-CS, though nursing facility discharge has been associated with increased long-term mortality in AMI-CS. Further research is needed to evaluate the impact of discharge disposition after a CS admission by sex [25–26].

### 6.2. Limitations

This study relies on the accuracy of administrative data to identify patients with cardiogenic shock. However, multiple contemporary CS registries have used a similar approach, and prior studies examining the accuracy of the R57.0 ICD-10 code to identify CS have reported its positive predictive value to be as high as 93% [27].

We also rely on administrative data to classify the different CS etiologies and collect their relevant management procedures and outcomes. The accuracy of these codes is less well-studied.

Also, complications were not assessed for patients treated with IABP or medical therapy alone. However, prior studies have shown that major complications in these patients have a low incidence in CS [28]. On the other hand, treatment with advanced MCS devices like VA-ECMO and Impella carry a higher complication rate, and their benefit is more contended among women [28–30].

Finally, our study did not collect data on readmissions or outcomes beyond initial discharge and hence we cannot assess the mid and long-term course of these patients.

### 6.3 Conclusion

This study provides a detailed overview of the sex-specific differences in nonAMI-CS, including distinct clinical profiles, etiologic variations, and management trends between men and women. Our findings underscore the importance of sex-based approaches in CS management and can help guide future research into strategies to improve care and optimize outcomes in this heterogeneous patient population.

## Data Availability

The raw data supporting the conclusions of this article will be made available by the authors upon request, without reservation.

## 7. Acknowledgments

None

## Non-standard Abbreviations and Acronyms

BARC: Bleeding Academic Research Consortium
CCI: Charlson Comorbidities Index
CSWG: Cardiogenic Shock Working Group
HF-CS: Heart failure related cardiogenic shock
NonAMI-CS: Non-acute myocardial infarction related cardiogenic shock
RRT: Renal replacement therapy
SCAI: Society for Cardiovascular Angiography and Interventions
SHARC: Shock Academic Research Consortium

## 10. Supplemental material

Supplemental table S1-S2 Supplemental Figures S1-S6

